# RAGCare-QA: A Benchmark Dataset for Evaluating Retrieval-Augmented Generation Pipelines in Theoretical Medical Knowledge

**DOI:** 10.1101/2025.08.15.25333718

**Authors:** Jovana Dobreva, Ivana Karasmanakis, Filip Ivanisevic, Tadej Horvat, Dimitar Kitanovski, Matjaz Gams, Kostadin Mishev, Monika Simjanoska Misheva

**Author notes:** **Corresponding author’s email address and Twitter handle**.

## Abstract

The paper introduces RAGCare-QA, an extensive dataset of 420 theoretical medical knowledge questions for assessing Retrieval-Augmented Generation (RAG) pipelines in medical education and evaluation settings. The dataset includes one-choice-only questions from six medical specialties (Cardiology, Endocrinology, Gastroenterology, Family Medicine, Oncology, and Neurology) with three levels of complexity (Basic, Intermediate, and Advanced). Each question is accompanied by the best fit of RAG implementation complexity level, such as Basic RAG (315 questions, 75.0%), Multi-vector RAG (82 questions, 19.5%), and Graph-enhanced RAG (23 questions, 5.5%). The questions emphasize theoretical medical knowledge on fundamental concepts, pathophysiology, diagnostic criteria, and treatment principles important in medical education. The dataset is a useful tool for the assessment of RAG-based medical education systems, allowing researchers to fine-tune retrieval methods for various categories of theoretical medical knowledge questions.

**VALUE OF THE DATA:**

– RAGCare-QA dataset is designed to benchmark state-of-the-art RAG architectures recommendations for theoretical medical knowledge through 420 human annotated single-choice questions, well-distributed in 6 different medical specialties.
– Researchers can leverage this resource to build more effective educational tools that adapt their retrieval strategies based on question complexity and medical specialty.
– The dataset fills a gap in medical AI by providing a standardized benchmark that supports the development of AI-based adaptive educational tools.
– The dataset classifies each question by the most suitable RAG architecture, Basic, Multi-vector, or Graph-enhanced, needed for context retrieval, enabling precise performance comparisons across retrieval strategies.
– The dataset can serve as a foundation for development of specialized retrieval strategies to enhance learning outcomes in medical education.

## BACKGROUND

The integration of artificial intelligence (AI) in medical education has gained significant momentum, with RAG systems showing particular promise for knowledge assessment and educational content delivery [1, 2]. While large language models (LLMs) demonstrate substantial medical knowledge, their performance in educational contexts is significantly enhanced when combined with specialized retrieval systems that access curated medical educational content [3, 4].

Theoretical medical knowledge assessment has particular requirements distinguishing it from clinical problem-solving exercise. Didactic questions require precise recall of bottom-line ideas, pathophysiologic processes, and traditional medical principles from structured sets of knowledge [5, 6]. The depth of medical theoretical knowledge, from straightforward definitions to intricate pathophysiological correlations, requires sophisticated retrieval mechanisms that can be calibrated for differing levels of cognition.

Existing medical QA datasets, including PubMedQA [5], MedMCQA [6], and specialized disease-focused collections [7], primarily emphasizing clinical decision-making rather than systematic evaluation of retrieval pipelines for educational content. These datasets evaluate model performance against established medical knowledge, however, do not address how different RAG pipelines influence learning and assessment outcomes in educational settings. The landscape of RAG pipelines offers multiple approaches, each with distinct advantages for educational applications.

Basic RAG implementations provide straightforward document retrieval suitable for direct factual queries common in foundational medical education [8]. Multi-vector RAG models show promise in managing various types of educational content, ranging from dictionary-style definitions to elaborate explanations, making them a good fit for inclusive medical education applications [9, 10]. Graph-augmented RAG systems excel at representing hierarchical medical knowledge structures and complex concept relationships essential for advanced theoretical understanding [11, 12]. These pipeline designs have shown to be useful in educational settings where information must be accessed from several angles and sources of knowledge.

There is a notable gap in the medical AI community regarding systematic approaches to choosing suitable RAG pipelines for different categories of theoretical medical knowledge questions, which often leads to suboptimal design of educational systems. Although recent advances, such as HuatuoGPT

[13] and other education-oriented AI models, have contributed to medical education, they have largely prioritized architectural improvements over the optimization of retrieval strategies for instructional content.

This dataset bridges this important gap by offering a systematically annotated set of theoretical medical questions with specific RAG pipeline recommendations, allowing evidence-based retrieval strategy selection for medical education use cases.

## DATA DESCRIPTION

### Dataset Structure and Composition

The RAGCare-QA dataset comprises 420 theoretical medical knowledge questions systematically distributed across medical specialties, complexity levels, and RAG implementation categories. Table 1 provides a detailed breakdown of the dataset structure.

**Table 1.** Dataset Breakdown by Medical Specialty and Complexity Levels.

| Medical Specialty | Basic | Intermediate | Advanced | Total |
| --- | --- | --- | --- | --- |
| Cardiology | 32 | 36 | 19 | 87 |
| Endocrinology | 28 | 32 | 14 | 74 |
| Family Medicine | 26 | 41 | 14 | 81 |
| Gastroenterology | 20 | 22 | 12 | 54 |
| Neurology | 20 | 22 | 15 | 57 |
| Oncology | 24 | 28 | 15 | 67 |
| **Total** | **150** | **181** | **89** | **420** |

Complexity Level Distribution:

- Basic (150 questions): Fundamental medical concepts, definitions, and straightforward factual knowledge.
- Intermediate (181 questions): Moderate complexity questions involving pathophysiology, diagnostic criteria, and treatment principles.
- Advanced (89 questions): Complex theoretical scenarios requiring deep understanding of medical mechanisms and advanced concepts.

RAG Implementation Complexity Distribution:

- Basic RAG: 315 questions (75.0%) - Direct factual queries with explicit terminology and straightforward retrieval requirements.
- Multi-vector RAG: 82 questions (19.5%) - Questions requiring diverse knowledge sources and multiple representation approaches.
- Graph-enhanced RAG: 23 questions (5.5%) - Complex relationship-based queries requiring structured knowledge representation.

Each entry in the dataset follows a structured format with the following fields:

- Type: Medical specialty classification.
- Question: Multiple-choice question with options (a-e).
- Answer: Correct answer designation (a, b, c, d, or e).
- Text Answer: Correct answer in textual format
- Reference: Citation of the medical literature source.
- Page: Specific page reference within the source.
- Context: Relevant text from source that supports the answer.
- Label: Optimal RAG pipeline classification.
- Complexity: Difficulty level based on the depth of medical knowledge and clinical reasoning required to answer the question

### Dataset Statistics and Characteristics

The RAGCare-QA dataset exhibits diverse features across question complexity and content structure. Table 2 provides detailed statistical analysis of the dataset composition.

**Table 2.** RAGCare-QA Statistical Features.

| Feature | Minimum | Average | Maximum |
| --- | --- | --- | --- |
| Question Length (words) | 12 | 47 | 112 |
| Answer Options | 5 | 5 | 5 |
| Context Length (words) | 25 | 89 | 245 |
| Publication Years | 1985 | 2015 | 2024 |
| **Reference Types:** Medical Textbooks (63%), Peer-reviewed Journals (30%), Other Sources (7%) |  |  |  |

#### Answer Distribution Analysis

The correct answers are well-distributed across all five options, ensuring balanced assessment: Option A (22%), Option B (20%), Option C (19%), Option D (21%), Option E (18%). This distribution prevents systematic bias toward specific answer positions. Source Reference Analysis: Reference Source Distribution: The dataset incorporates a balanced mix of authoritative medical sources: medical textbooks (63%), peer-reviewed journal articles (30%), and other specialized medical resources (7%). The primary sources include “Interna medicina” (6th edition, 2022) [14] and “Harrison’s Principles of Internal Medicine” [15] for foundational knowledge, supplemented by high-impact journal publications. Publication years span from 1985 to 2024, ensuring coverage of both foundational medical knowledge and current research developments.

Complexity Progression: Questions demonstrate clear complexity escalation from Basic (average 32 words) to Intermediate (average 48 words) to Advanced (average 65 words), reflecting increasing cognitive load and knowledge integration requirements

### Question Examples by RAG Pipeline Type

The dataset encompasses various types of theoretical medical knowledge questions, each designed to test specific aspects of medical understanding, with optimal RAG pipeline assignments based on the framework shown in Figure 1.

**Figure 1.**
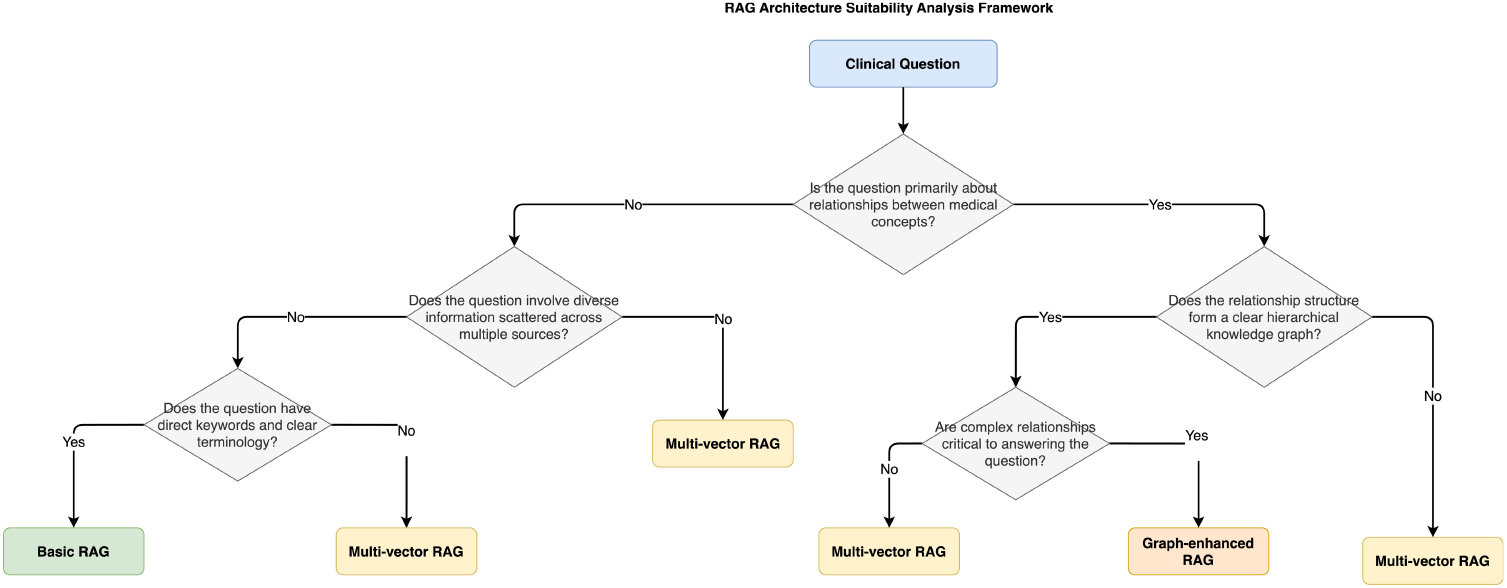
RAG Pipeline Classification Decision Tree. The figure presents the systematic decision-making framework used to classify theoretical medical questions into their optimal RAG pipeline categories. The decision tree evaluates key characteristics such as relationship complexity between medical concepts, information distribution patterns, query structure complexity, and reasoning requirements. Starting from the initial question analysis, the framework guides classification into Basic RAG (for direct factual queries), Multi-vector RAG (for questions requiring diverse knowledge sources), or Graph-enhanced RAG (for complex relationship-based queries). This evidence-based classification ensures optimal matching between question types and retrieval pipelines for medical education applications.

### Complexity Level Characteristics

The questions in the dataset are categorized into three complexity levels: Basic, Intermediate, and Advanced based on the depth of medical knowledge required, the cognitive effort involved, and the nature of information integration needed for accurate resolution:

- Basic Level Questions focus on fundamental definitions, basic pathophysiology, and direct factual knowledge. These questions typically involve single-concept retrieval and straightforward medical terminology recognition.
- Intermediate Level Questions involve moderate complexity scenarios requiring understanding of disease mechanisms, diagnostic approaches, and treatment principles. These questions often require integration of multiple medical concepts and represent the largest portion of the dataset.
- Advanced Level Questions present complex theoretical scenarios demanding deep understanding of pathophysiological mechanisms, differential diagnosis considerations, and advanced medical principles. These questions often involve sophisticated medical reasoning and comprehensive knowledge integration.

### RAG Pipeline Classification Framework

Each question underwent systematic analysis to determine its optimal RAG pipeline suitability using a structured evaluation framework. The decision-making process follows a systematic classification tree as illustrated in Figure 1, while the resulting pipeline types are compared in Figure 2. The classification framework evaluates several key factors:

**Figure 2.**
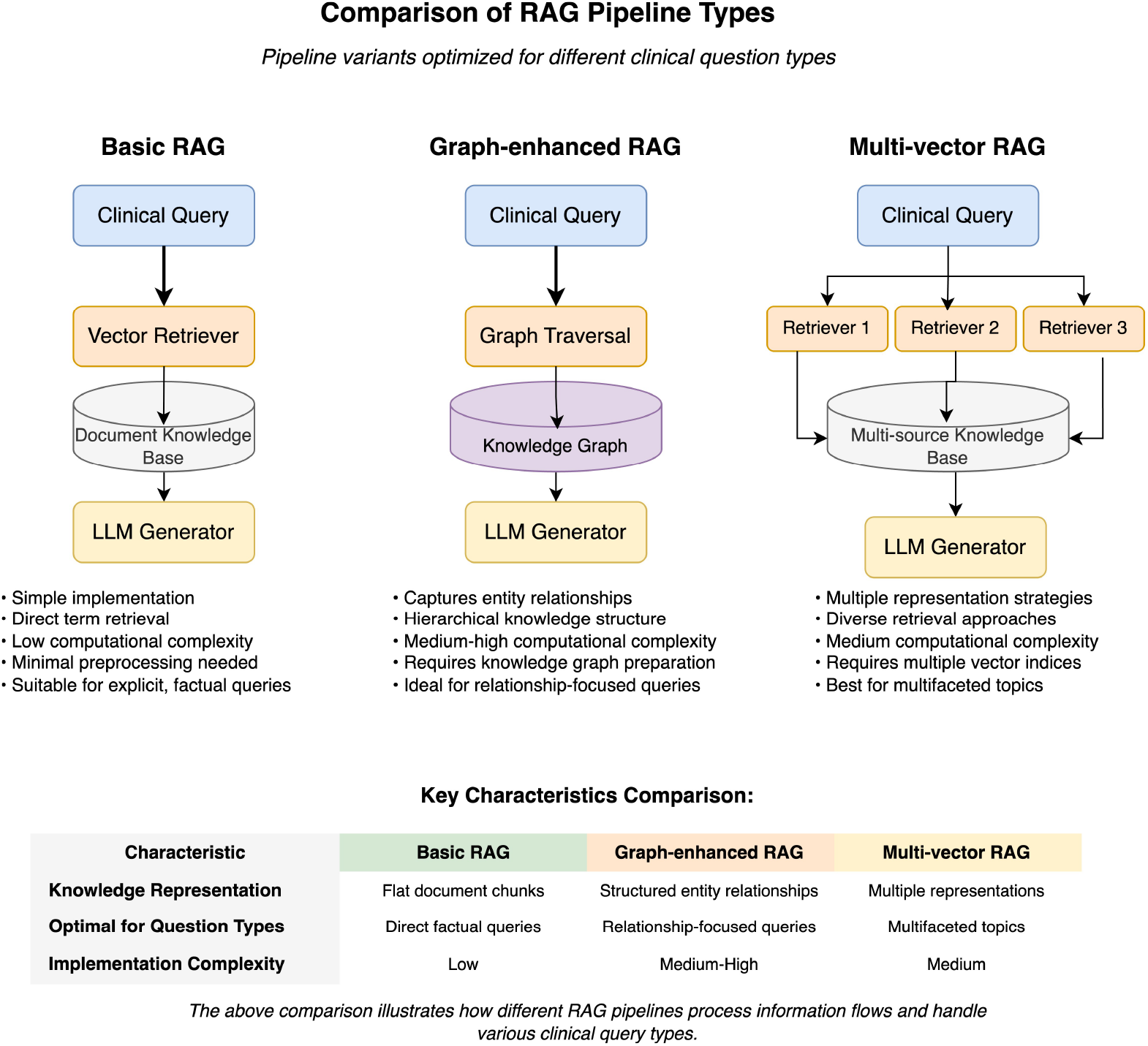
Comparison of RAG Pipeline Types. The figure illustrates the key differences between Basic RAG (simple document retrieval), Graph-enhanced RAG (structured knowledge representation), and Multi-vector RAG (diverse representation approaches). For each pipeline type, the diagram shows information flow patterns, typical use cases, and relative computational complexity. The comparison table highlights how different medical question types benefit from specific design approaches optimized for theoretical medical knowledge retrieval.

Basic RAG Suitability (78.75% of questions): Questions with explicit terminology, direct factual content, and straightforward retrieval requirements. These typically involve definition-based queries or simple factual recall that can be effectively answered through standard document retrieval. Multi-vector RAG Suitability (20.5% of questions): Questions requiring diverse knowledge sources, multiple perspectives on medical concepts, or integration of information from various medical domains. These questions benefit from multiple representation strategies and comprehensive knowledge coverage.

Graph-enhanced RAG Suitability (5.5% of questions): Questions involving complex medical relationships, hierarchical knowledge structures, and entity interconnections that benefit from graph-based knowledge representation. These represent sophisticated theoretical queries requiring advanced reasoning capabilities.

By systematically considering information complexity, retrieval needs, domain breadth, and cognitive demands, the classification process ensures each question is aligned with the most suitable RAG pipeline to maximize performance in educational applications.

## EXPERIMENTAL DESIGN, MATERIALS AND METHODS

### Dataset Creation Process

The RAGCare-QA dataset was developed through a systematic multi-stage process specifically designed for theoretical medical knowledge assessment, as illustrated in Figure 3. The creation process began with comprehensive collection of source materials from authoritative European medical textbooks, educational curricula, and clinical guidelines commonly used in medical education programs.

**Figure 3.**
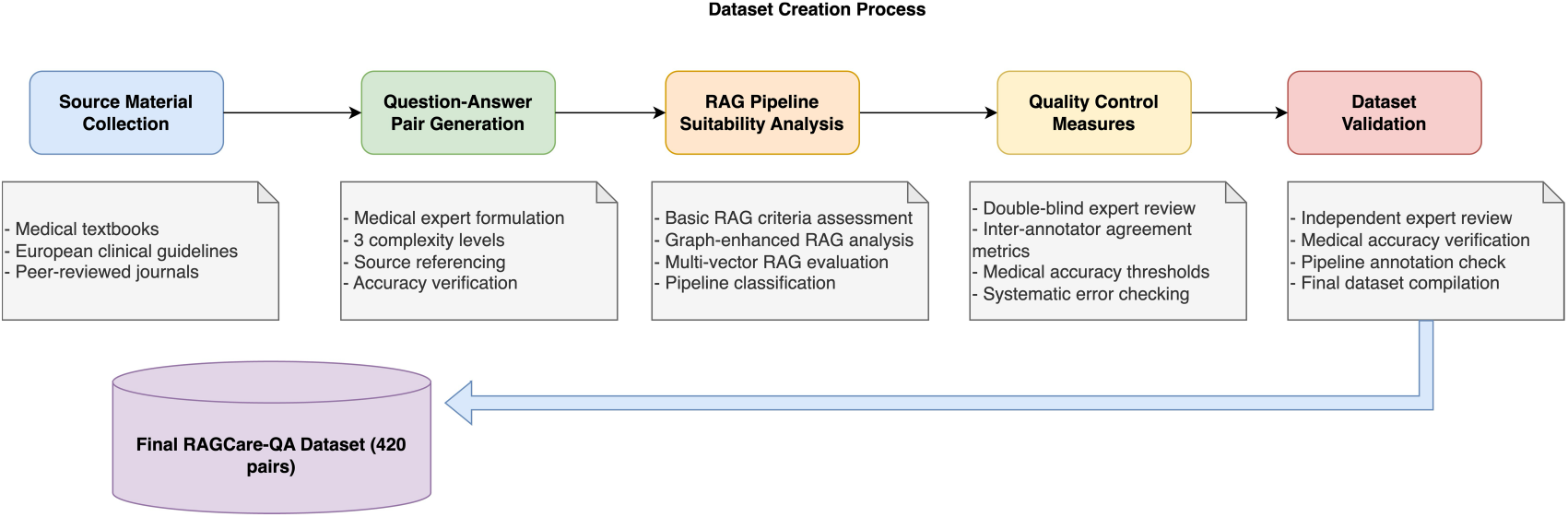
Dataset Creation Process. The figure illustrates the systematic five-stage approach used to develop the RAGCare-QA dataset. The process begins with source material collection from European medical literature, followed by question-answer pair generation by medical experts across six specialties and three complexity levels. Each pair then undergoes RAG pipeline suitability analysis using specific criteria for Basic RAG, Graph-enhanced RAG, and Multi-vector RAG approaches. Quality control measures ensure medical accuracy and appropriate complexity classification. The process concludes with dataset validation by independent experts. The final dataset contains 420 theoretical medical knowledge questions with comprehensive annotations and source references.

The source materials were systematically collected from authoritative medical references across multiple licensing categories to ensure comprehensive coverage and copyright compliance. The primary source was “Interna medicina” (6th edition, 2022), editors Mitja Košnik, Dušan Štajer, and colleagues, published by Medicinska fakulteta (University of Ljubljana Medical Faculty), Slovensko zdravniško društvo (Slovenian Medical Association), and Buča [14], contributing 180 questions (42.9% of the dataset). Additional sources included “Onkologija: Učbenik za študente medicine” (1st edition, 2018) by Strojan & Hočevar, published by Onkološki inštitut Ljubljana [16], contributing 44 questions (10.5%) and licensed under Creative Commons Attribution-NonCommercial-NoDerivatives 4.0 International. “Harrison’s Principles of Internal Medicine” (20th edition) [15] contributed 19 questions (4.5%) through transformed contextual synthesis. Open access educational resources from StatPearls and NCBI Bookshelf [17] provided 56 questions (13.3%), also licensed under Creative Commons Attribution-NonCommercial-NoDerivatives 4.0 International. Peer-reviewed journal articles from high-impact publications contributed 39 questions (9.3%), and the remaining 82 questions (19.5%) were derived from additional educational materials including medical curricula and clinical practice guidelines. This multi-tiered approach ensures comprehensive coverage of established medical knowledge across six targeted specialties while maintaining rigorous copyright compliance standards.

Medical education experts and subject matter specialists formulated questions across six medical specialties, ensuring coverage of fundamental theoretical concepts essential for medical training. Questions were designed as multiple-choice items with five options (a-e), following standard medical education assessment formats. Each question targets specific learning objectives and cognitive levels appropriate for medical students and healthcare professionals.

The question development process prioritized theoretical knowledge over clinical case-based scenarios, focusing on pathophysiology, disease mechanisms, diagnostic criteria, pharmacological principles, and anatomical knowledge. This approach ensures the dataset serves as a comprehensive resource for foundational medical education rather than clinical decision-making training.

For each question, detailed contextual information was extracted from source materials, including specific page references and relevant text passages that support the correct answer. This contextualization enables effective training and evaluation of RAG pipelines by providing rich source material for retrieval processes.

### RAG Pipeline Analysis and Annotation

Each question underwent systematic analysis to determine its optimal RAG pipeline using a structured evaluation framework. The analysis considered multiple factors including information structure, retrieval complexity, knowledge domain requirements, and cognitive processing needs.

Basic RAG Classification: Questions were classified as Basic RAG suitable when they involved direct factual retrieval, explicit terminology, clear question structure, and straightforward answer pathways. These questions typically require simple document-based retrieval without complex relationship processing and represent the majority of the dataset (78.75%).

Multi-vector RAG Classification: Questions requiring diverse information sources, multiple conceptual perspectives, cross-domain knowledge integration, or comprehensive coverage of medical topics were classified as Multi-vector RAG suitable (20.5%). These questions benefit from multiple retrieval strategies and diverse representation approaches.

Graph-enhanced RAG Classification: A meaningful subset of questions (5.5%) requiring complex relationship modeling, hierarchical knowledge representation, and sophisticated entity interconnection analysis were classified as Graph-enhanced RAG suitable. These questions involve the most complex theoretical reasoning scenarios requiring advanced graph-based retrieval approaches.

The annotation process involved medical education experts working in conjunction with AI specialists to ensure both medical accuracy and appropriate technical classification using the decision framework shown in Figure 1. Inter-rater reliability was maintained through systematic review processes and consensus-building approaches.

### Quality Assurance and Validation

The dataset underwent comprehensive quality assurance to ensure medical accuracy, appropriate difficulty progression, and correct RAG pipeline classification. Medical content was validated against authoritative sources, with particular attention to European medical practice standards and educational requirements.

Each question’s difficulty level was validated through expert review, ensuring appropriate classification into Basic, Intermediate, and Advanced categories. The progression from basic factual knowledge to complex theoretical understanding reflects authentic medical education pathways.

RAG pipeline annotations were validated through cross-verification processes, where three experts independently assessed question suitability for different retrieval approaches. Disagreements were resolved through consensus meetings and detailed discussion of classification criteria. The final dataset underwent validation by independent medical and AI experts to ensure accuracy of medical content and appropriate RAG pipeline annotations, as shown in the final stage of Figure 3.

## LIMITATIONS

The RAGCare-QA dataset has several limitations that researchers should consider. The content primarily reflects European medical education standards and may not fully represent global medical education approaches or regional variations in medical practice. The dataset focuses on six major medical specialties, potentially limiting applicability to other medical domains such as surgery, pediatrics, or psychiatry.

The multiple-choice format, while standard in medical education, may not capture all forms of theoretical medical knowledge assessment used in educational settings. The dataset’s theoretical focus excludes practical clinical skills, procedural knowledge, and patient interaction scenarios that form important components of comprehensive medical education.

The complexity level classifications, while expert-validated, may not align perfectly with all educational frameworks or institutional standards. The RAG implementation complexity shows a reasonable balance among Basic RAG (75.0%) for straightforward queries, Multi-vector RAG (19.5%) for complex knowledge integration, and Graph-enhanced RAG (5.5%) for relationship-based reasoning, which may provide sufficient diversity for comprehensive retrieval architecture evaluation.

Language limitations exist as the dataset is primarily in English with some source materials in Slovenian, potentially affecting applicability in multilingual educational contexts. The static nature of the knowledge base may require periodic updates to maintain currency with evolving medical understanding and educational standards.

## Data Availability

All data produced are available online at https://huggingface.co/datasets/ChatMED-Project/RAGCare-QA

https://huggingface.co/datasets/ChatMED-Project/RAGCare-QA

## ETHICS STATEMENT

This research complies with ethical publication guidelines and institutional review requirements. The dataset construction involved no human subjects research, animal experimentation, or collection of sensitive personal data. All source materials were derived from publicly available medical literature, educational resources, and established clinical guidelines.

### Source Material Usage and Copyright Compliance

The dataset creation process employed systematic content transformation methodologies to ensure copyright compliance while maintaining educational research integrity. Question development involved creating original multiple-choice assessments based on established medical knowledge, with all contextual information paraphrased and synthesized from source materials rather than reproduced verbatim.

For materials licensed under Creative Commons Attribution-NonCommercial-NoDerivatives 4.0 International, including “Onkologija: Učbenik za študente medicine” [16] (44 questions, 10.5%) and StatPearls/NCBI Bookshelf resources [17] (56 questions, 13.3%), educational use is explicitly permitted with proper attribution. For “Interna medicina” [14] (180 questions, 42.9%), published by University of Ljubljana Medical Faculty in collaboration with Slovenian Medical Association, question development followed academic fair use principles for educational research involving university-published materials. For “Harrison’s Principles of Internal Medicine” [15] (19 questions, 4.5%), contextual information was substantially paraphrased and synthesized to present medical concepts in original formulations, preserving only bibliographic references and page citations for academic attribution purposes.

The systematic content paraphrasing process involved medical experts reformulating all medical concepts into novel assessment formats without reproducing any substantial portions of original textual content from any source. Only essential bibliographic information (references and page numbers) was preserved to maintain academic integrity and enable verification of medical accuracy. This comprehensive paraphrasing methodology represents original scholarly work that enhances medical education research while respecting intellectual property rights through complete textual transformation of all source materials.

No proprietary or confidential medical information was included in the dataset. The questions and answers represent established medical knowledge reformulated into original assessment items and do not include experimental or unvalidated medical information.

## CRediT AUTHOR STATEMENT

Jovana Dobreva: Conceptualization, Data curation, Investigation, Methodology, Writing - Original draft, Writing - Review & editing. Ivana Karasmanakis: Medical validation, Resources. Filip Ivanisevic: Medical validation, Resources. Tadej Horvat: Data curation, Formal analysis, Software, Validation. & editing. Dimitar Kitanovski: Investigation, Resources, Validation. & editing. Matjaz Gams: Supervision. Kostadin Mishev: Supervision, Formal analysis, Methodology, Writing - Review & editing. Monika Simjanoska Misheva: Conceptualization, Project administration, Supervision, Writing - Review & editing.

## ACKNOWLEDGEMENTS

Views and opinions expressed are, however, those of the author(s) only and do not necessarily reflect those of the European Union or the European Research Executive Agency. Neither the European Union nor the granting authority can be held responsible for them.

Funded by the European Union under Horizon Europe project ChatMED - Bridging Research Institutions to Catalyze Generative AI Adoption by the Health Sector in the Widening Countries (grant agreement ID: 101159214).

## DECLARATION OF COMPETING INTERESTS

The authors declare no competing financial interests or personal relationships that could influence the work reported in this paper.

## Notes

### Competing Interest Statement

The authors have declared no competing interest.

### Funding Statement

This study was funded by the European Union under Horizon Europe project ChatMED - Bridging Research Institutions to Catalyze Generative AI Adoption by the Health Sector in the Widening Countries (grant agreement ID: 101159214).

